# Identification of Cognitive Impairment in Cardiovascular Rehabilitation: A Cross-Sectional Study Protocol

**DOI:** 10.1101/2021.01.01.21249114

**Authors:** Qutub Jamali, Salman Karim, Mirza Najiullah Beg, Muhammad Shoaib Raja, Kalpesh Solanki, Chukwuma Oraegbunam

## Abstract

**Introduction:** Cardiac Rehabilitation is a multidisciplinary intervention for people after an adverse cardiac event to improve their physical, psychological and social functioning. The risk factors of cardiac disease and dementia are similar. This cross sectional study aims to determine whether adding memory assessment to a cardiac rehabilitation program would improve early detection of cognitive impairment.

**Methods and Analysis:** Participants will undergo cognitive assessment by using Addenbrooke’s Cognitive Examination (ACE-III). The data obtained will be divided into: - 1- Participants who had a history of memory problems before and after the adverse cardiac event. 2-Participants with no history of memory problems before and currently presents with cognitive impairment. 3-Participants with no memory problems before and after the adverse cardiac event.

**Ethics and dissemination:** Study ethical approval has been granted by Sheffield Research Ethics Committee (reference 20/YH/0146) and the NHS Health Research Authority (project reference 273763).

## Introduction

Dementia is a progressive, irreversible disorder of the brain causing focal cognitive deficit affecting the activities of daily living. When a person is diagnosed with Dementia, it not only affects the individuals themselves but also their family. According to a report from the Alzheimer’s Society in 2016, there are a number of cardiovascular risk factors associated with dementia including type-2 diabetes in mid-life or later life, high blood pressure in mid-life, high total blood cholesterol levels in mid-life and obesity in mid-life. Having cardiovascular disease or type-2 diabetes mellitus doubles a person’s risk of developing dementia^2^□.

In the early stages of Dementia, denial of changes in cognition, functional ability, mood or behaviour are common coping strategies. This can lead to family conflict. For example, when a person has been brought to see the GP by a concerned family member, the person can become angry and may feel betrayed. As the person’s denial strengthens, it can cause significant carer stress. On other occasions, both family member and the affected person make adjustments and mask the memory problem by acting as the memory for the other^2^□. This leads to delay in diagnosis of Dementia. Timely detection and diagnosis will prevent crisis, facilitate adjustment and provide access to treatments and support^2^□. Therefore, an early diagnosis could be beneficial for the patient and their family as they would be able to access support and care appropriately. This would help with plans for future decision making, engagement with community support, understanding of the illness, symptoms and plans for future care.

Cardiac Rehabilitation (CR) is a multidisciplinary intervention for people after myocardial infarction (MI), heart failure or post cardiac surgery, to improve their physical, psychological, social and vocational functioning. It is an integrated approach comprising of around 10 to 12 weeks of structured physical exercise, behavioural change aimed at healthier lifestyle, risk factor management, group health education sessions and psychological interventions^1^□^2^. CR reduces mortality and morbidity following an adverse cardiac event and improves the quality of life^3^□^7^.

It has been found that people suffering from heart failure (HF) are at a greater risk of developing Alzheimer’s or Vascular Dementia^9^. Morris et al. in 2019 suggested that chronic obstructive pulmonary disease (COPD) is a major risk factor for cognitive impairment^24^. A study conducted by Barclay et al in 1988 indicated that mild cognitive impairment may be under-diagnosed in CR^10^. A long term study exploring a 12 month follow up CR program by Alosco et al in 2014 showed significant improvements in memory and cognitive function^13^.

CR has proved to be effective in improving the memory and executive functions of patients who undergo a structured exercise program^11^□ ^12^. People with mild cognitive impairment who are at a potential risk of developing Dementia have benefitted from a progressive supervised exercise program^14^□^17^. CR can be a preventive measure against Dementia^1^□□^19^. It adds further value considering the fact that the average age of a person participating in CR in the UK in 2018 was 67 years^8^ which is an ideal age for the assessment of cognitive function. Early detection of dementia is the key and cardiac rehabilitation could potentially be the best possible solution to address the issue as cardiac co-morbidities are one of the major risk factors for dementia.

A qualitative study conducted by Sunball et al. in 2018 examined the long term risk of dementia after first MI. They found that heart attack increased the risk of vascular dementia by 35% and the risk remains higher for up to 35 years after the heart attack^2^5 which clearly justifies the importance of early diagnosis.

Most of the studies done in the past have emphasized mild cognitive impairment. A recent study by Mohammad et al. in 2019 emphasized that people participating in CR can potentially have poor memory and poor executive function^22^. However, the possibility of false positives was high because the screening tool used was Mini-MOCA which is very brief.

Depression, anxiety and fear of recurrence after myocardial infarction often cause disability which could be more severe than the actual cardiac impairment. As far back as 1977 Soloff had recommended that a liaison psychiatrist can play a crucial role in cardiac rehabilitation through individual consultation and team meetings to address the psychosocial elements of the condition^20^. However, despite several studies highlighting the benefits of CR in improving mental health and overall improvement in quality of life, the general perception is that Psychiatry involvement is considered as a complementary component rather than essential in a CR program^21^. The overall involvement of Psychologist in 2018 was 24% and Doctor (Specialist unspecified) was 10% of total CR centres in the UK^12^.

A study by Intzandt et al. in 2015 emphasized on the feasibility of referring patients with MCI in CR^23^. However, the quality of data was limited as it was relying more on self-reports and a longitudinal trial was suggested. A Randomised Controlled Trial conducted by McGuinness et al. in 2015 showed that in people with mild cognitive impairment, the average time span of progression to Dementia is around 30 months (2.5 years) ^2^□. Moreover, another study by Farias et al. in 2009 report that the time it takes to progress from mild cognitive impairment to dementia is 2.4 years if it is associated with risk factors such as hypertension, diabetes and heart disease^30^.

### Objectives/Research Question

Currently, cognitive assessment is not included in the core components of a cardiac rehabilitation program. The aim of this study is to determine whether adding memory assessment to a cardiac rehabilitation program would improve early detection of cognitive impairment.

Therefore, the research question is:-

**Can adding a memory assessment to the cardiac rehabilitation program improve early detection of cognitive impairment in people with cardiac disease?**

## Methods and Analysis

### Inclusion criteria

- Male and female.
- Age ≥ 40 years. There is no upper age limit.
- Patients attending cardiac rehabilitation in phase 2/3/4.
- People attending CR unit at UHMB NHS Trust.
- English speaking people.
- People with heart attack within the last six months.
- Heart failure.
- Post CABG/PTCA/Valve Replacement.
- People with a past history of heart attack/stable angina who are currently undergoing phase-4 (maintenance) cardiac rehabilitation.
- Adults who have capacity to make decisions about their care and treatment.

Exclusion Criteria: -

- Age < 40 years
- People not attending CR Unit at UHMB NHS Trust.
- Non English speaking people.
- Very low mood, distressed and anxious.
- Adults who lack capacity to make decisions about their care and treatment.

### Design–

Firstly, participants will be recruited from cardiac rehabilitation department at University Hospitals of Morecambe Bay (UHMB) NHS Trust. Their first point of contact would be the cardiac rehab nurse/exercise physiologist/occupational therapist when they will come for their functional assessment for their cardiac problem before starting the exercise program which is the usual protocol in cardiac rehabilitation. At the end of their functional assessment, the cardiac rehab staff will ask whether the participant would be interested in going for a memory assessment. If the participant would like to know more about the study, they will be referred to the principal investigator. The principal investigator will then explain the study in detail, provide the participant information sheet and obtain consent from the participants.

If the participant has agreed for memory assessment, the principal investigator will meet the participant again for a detailed memory assessment which will include medical, psychiatric, cognitive function history (as mentioned in the patient demographic information sheet) and completing a memory assessment tool called Addenbrooke’s Cognitive Examination – ACE-III. However, if the principal investigator feels that the participant is not ready for the assessment which could be due to physical health issues or functional mental health problems like low mood, anxiety, then the assessment would not be conducted at that time. Since the duration of cardiac rehabilitation program is for 10 to 12 weeks, the assessment will be re-arranged when the participant is physically and mentally stable. The ACE-III score obtained from the participants will be recorded by the Principal Investigator.

The memory assessment can also be conducted at the participant’s home or at the memory assessment clinic at Altham Meadows, Morecambe, according to the convenience of the participants. It would be encouraged that a family member is also present during the assessment. Based on the history and ACE-III score, the principal investigator will send a referral to the Memory assessment Service according to the area of residence of the participant for further assessment and investigations, provided the participant has given his consent. Moreover, the principal investigator with the consent of the participant, will also send a letter to the participant’s GP mentioning about the assessment and referral.

The data will be collected in a paper copy which will be anonymised and stored securely. An Audit would be done twice, 1-After six months of the study and 2-At the end of the study to ensure that the data is collected according to the inclusion and exclusion criteria and is anonymised.

#### Materials

Questionnaire – Addenbrooke’s Cognitive Examination (ACE-III).

#### Sample Size

The duration of the study is one year and since it is a pilot study, we aim to recruit 200 participants in one year.

### Analysis

It will be a Cross-Sectional study to assess the prevalence of cognitive impairment in patients undergoing cardiac rehabilitation. The data obtained will be divided into three categories: -

1. Number of participants who had a history of memory problems before the adverse cardiac event (MI-heart attack and revascularization) and continue to have memory problems after the adverse cardiac event.
2. Number of participants with no history of memory problems before the adverse cardiac event and currently presents with cognitive impairment after the adverse cardiac event.
3. Number of participants with no memory problems before and after the adverse cardiac event.

## Ethics & dissemination

Study ethical approval has been granted by Yorkshire & The Humber - Sheffield Research Ethics Committee (reference 20/YH/0146) and the NHS Health Research Authority (project reference 273763). The Lancashire & South Cumbria NHS Foundation Trust accepted the role and responsibilities of study sponsorship. Protocol amendments approved by the Research Ethics Committee/NHS Health Research Authority will be communicated with the study sponsor, study team and where necessary, the trial registry.

The study aims to publish the findings in a peer-reviewed journal and present the results at National and International conferences.

## Discussion

This Study’s strength includes the use of a validated questionnaire for cognitive assessment which is ACE-III. If cognitive impairment is detected and treated early, it can either cure (if the cause is reversible) or can delay the progression of cognitive impairment (if the cause is irreversible). Moreover, along with the cross sectional data of detection of cognitive impairment, the assessment will include a detailed cognitive history using patient demographic information sheet (Appendix-1). This will provide very useful information such as the age of onset of memory problems, associated co-morbidities and lifestyle habits i.e. Cigarette smoking, tobacco or alcohol consumption.

The limitation of this study is that the patho-physiological changes associated with recent heart attack (Acute Myocardial Infarction) in a previously healthy individual are bound to be different from someone being rehabilitated after a diagnosis of heart failure, CABG/PTCA/Valve replacement. Moreover the patients with these conditions are more likely to have a history of Cardiac conditions for varying periods and with associated psychological impact/life changing adjustments. These are more than likely to impact their cognitive function.

It would be ideal to do a longitudinal study to determine whether a structured cardiac rehabilitation program would delay or slow the progression of cognitive impairment. Since it is a pilot study with no funding, the study will focus only on cognitive screening in cardiac rehabilitation.

## Trial Status

The study was approved by the Research Ethics Committee and NHS Health Research Authority in August 2020. Due to the ongoing COVID-19 pandemic, data collection process has been delayed. Participant recruitment and data collection will begin once the lockdown restrictions are eased.

## Data Availability

Currently its a study protocol. Data collection has not commenced yet.

## Appendix-I

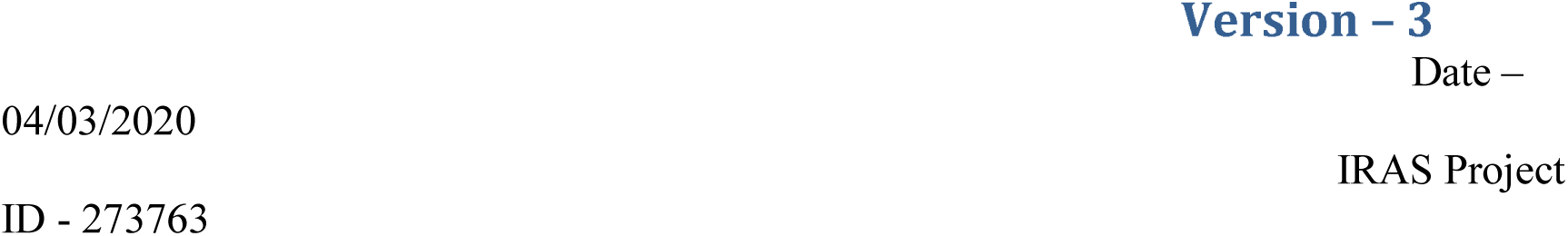

### Identification of Memory loss in Cardiovascular Rehabilitation

1- Screening Number –
2- Patient Name –
3- Date of Visit –
4- Age –
5- Sex –
6- Ethnicity –
7- List of Staff Present –
8- Medical history –

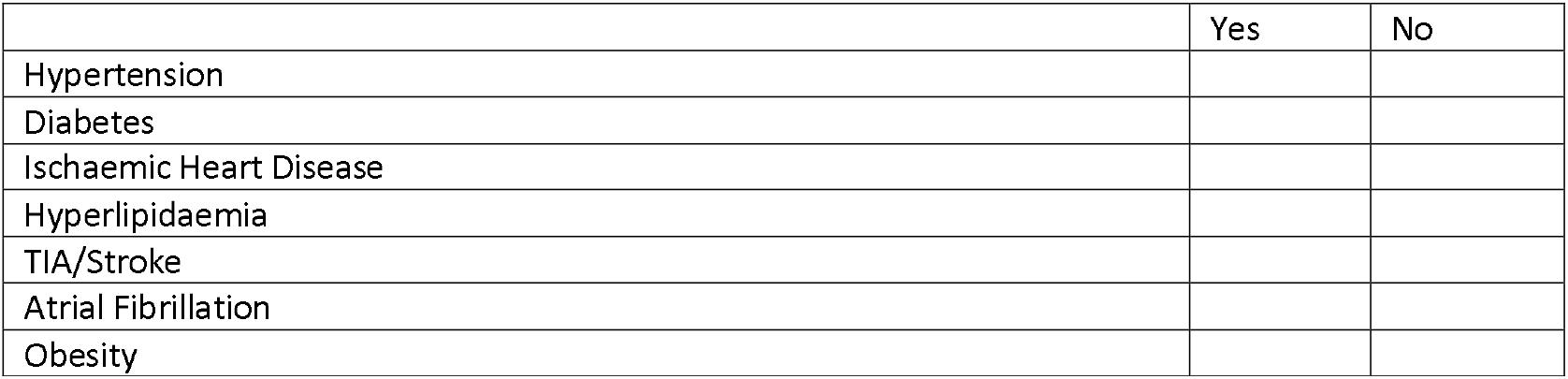
9- Past Psychiatric Study: -

|  | Yes | No |
| --- | --- | --- |
| Hypertension |  |  |
| Diabetes |  |  |
| Ischaemic Heart Disease |  |  |
| Hyperlipidaemia |  |  |
| TIA/Stroke |  |  |
| Atrial Fibrillation |  |  |
| Obesity |  |  |

10- History of Cognitive decline: - Duration: Nature: Impact on work and daily life: Issues of safety (for example, driving): Family history of dementia:
11- Alcohol use - Month and year of subject’s last use - How many years did you consume alcohol - Number of units consumed per day –
12- Tobacco use – Average number of cigarettes/cigars per day- How many years did you smoke-?
13- Cognitive function test: -

9- Past Psychiatric Study: -
|  | Yes | No |
| --- | --- | --- |
| Depression |  |  |
| Anxiety |  |  |
| Other: - |  |  |

14- Vital signs – Pulse- Blood pressure sitting – Blood pressure standing –
15- Weight –
16- Height –
17- BMI –

| **Patient** | **Completed**<br>**Y/N** | **Scale**<br>**completed by**<br>**(staff initial)** | **Date** |
| --- | --- | --- | --- |
| ACE-III |  |  |  |

